# Factors Influencing Integrated Disease Surveillance and Response System in selected Districts in the Eastern Region of Ghana

**DOI:** 10.1101/2023.12.05.23299500

**Authors:** Paul Twene, Bismark Sarfo, Alfred A. E Yawson, John Ekow Otoo, Annette Asraku

## Abstract

**Background:** Ghana adopted the Integrated Disease Surveillance and Response (IDSR) system, which is an integration of the various programs in the surveillance system and can contain disease outbreaks and pandemics. Implementation of the IDSR is influenced by several factors which can affect its functionality and ability to contain disease outbreaks. This study assessed the factors influencing the IDSR system in selected districts in the Eastern Region of Ghana.

**Method:** A cross-sectional study was conducted between February-March, 2022 in Fanteakwa North, Abuakwa South and New Juaben South districts in the Eastern Region of Ghana among health care workers who are involved in IDRS activities. Both primary and secondary data were collected and analyzed using descriptive statistics and regression analysis at 0.05 significant level with 95% confidence interval.

**Results:** Three hundred and forty-seven (347) health care workers participated in the study with 56.2% (195/347) indicating that rumor registers were available at the health facilities. Most of the respondents (64.8%, 225/347) had means of transport for disease surveillance activities while majority (61.9%, 215/347) had case-based forms for case investigation. About half (51.9%, 180/347) of the participants revealed that they did not receive any feedback from the next higher level in the past year. Availability of transport for IDSR activities was almost 3.4 times more likely to contribute positively to IDSR system compared to facilities without transport (AOR= 3.36; 95% CI= 1.44-7.83; p=0.005). Respondents who have the capacity to apply case definition are 2 times more likely to contribute to an effective IDSR system compared to health workers who cannot apply case definition (AOR= 1.94; 95% CI= 1.17-3.21; p=0.013). Respondents who did not receive feedback from the next higher level were 52% less likely to have an effective IDSR system compared to respondents who received feedback from the next higher level (AOR= 0.48; 95% CI= 0.23-1.00; p= 0.05).

**Conclusion:** Effective operation of IDSR is affected by the application of case definition and means of transport at health facilities. In addition, the capacity of health care workers to provide feedback can influence the smooth operation of the IDSR in the studied area in Ghana.

## Background

As part of efforts to improve upon public health surveillance and timely response for priority diseases conditions and events at all levels of service delivery, the Integrated Disease Surveillance and Response (IDSR) strategy was adopted by the World Health Organization in September 1998 [1]. Prior to 1998, the disease surveillance systems in most African countries were operating in a vertical manner where resources were used specifically for individual programs which led to some biases in the system [2]. As at December 2017, forty-four (44) countries in Africa representing 94% were implementing the IDSR strategy [3]. There were challenges identified with regards to the timeliness and completeness of reporting as only 32 (68%) of the countries achieved 80% of their reporting units [3]. The core functions of surveillance include case detection, reporting, investigation and confirmation, analysis and interpretation, response or action, feedback and evaluation.

Availability of trained human resource is paramount in surveillance activities and a functional disease surveillance system requires essential tools and job aids at all levels of service delivery which include the facility register, case investigation forms for disease-specific and generic, weekly and monthly reporting forms, and standard case definitions. In Southern Ethiopia, routine reporting is frequently of poor quality due to a variety of issues, including low motivation, inadequate supervision and feedback, and staff overburdening from multiple disease-specific reporting requests [4].

The resources needed for such routine activities in most cases do not have available funding for their implementations [5]. In the Eastern Region, the IDSR is well streamlined into the health care system. The structure of the surveillance system is divided into four levels; regional, district, sub-district and the Community-based Health Planning and Services (CHPS) level.

Ghana is faced with multiple barriers to IDSR implementation due to limited resources and weak healthcare infrastructure [6]. In the Eastern Region, IDSR system is challenged with multiple factors in varied forms. At the Municipal and District levels, routine surveillance is challenged with inadequate number of trained personnel, inadequate laboratory facilities, financial resource constraints, lack of transport systems, and late and inadequate reporting of surveillance data[6].

The core surveillance functions are key to the implementation of an ideal disease surveillance system however, there is limited information pertaining to the integration of the various aspects of IDSR in Ghana and for that matter, the Eastern Region. Findings from this study would be useful in strengthening the implementation of IDSR system focusing on the core diseases surveillance functions in the districts. We hope that this study provides information to help strengthen the disease surveillance system and better position the region to detect and prevent disease outbreaks.

## Methods

### Setting of the Study

The study was carried out in the health facilities in two municipals, namely New Juaben South and Abuakwa South and one district which was Fanteakwa North.

### Abuakwa South

The Abuakwa South Municipality is one of the 33 administrative Districts in the Eastern Region of Ghana. The municipality has a projected population of 10,0830 for year 2021[7]. The municipality has 1 Hospital, 1 Clinic, 4 Health Centres, 15 CHPS compounds and demarcated CHPS Zones. The district operates both active and passive surveillance system on routine basis with health professionals and a network of Community Based Surveillance Volunteers. The municipal Health Directorate has a total of 185 health care workers across the fifty (50) health facilities.

### New Juaben South

New Juaben south is one of the municipalities, and has an estimated total population of 156, 879 in 2021 with eighty-five (85) communities. The major referral health facility, Regional Hospital is located within the municipality which is supported by other health facilities which include, 12 CHAG Hospitals, 5 Health Centres, 11 Private Clinics, 1 Polyclinic1, and 35 CHPS compounds, a total of 51 health facilities [8].The municipal Health Directorate has a total of 2,297 health care workers.

### Fanteakwa North

Fanteakwa North district has a projected population of 76,434 for the year 2021.The health delivery system in the district is carried out by staff working in thirty-one (31) public and private health institutions with staff strength of three hundred and forty five (345) health workers [9]. These institutions are all government establishment except Salvation Army clinic which is a member of the Christian Health Association of Ghana (CHAG) institution. The strongest strength of the district in terms of community health work is the Community-based Surveillance programme. One hundred and fifty (150) functional and active Community Based Surveillance Volunteers (CBSVs) have been trained to support community health activities. They record and report on epidemic-prone diseases, deliveries and deaths in their catchment areas on monthly basis and also support the health directorate during mass campaigns [9].

**Fig 1:**
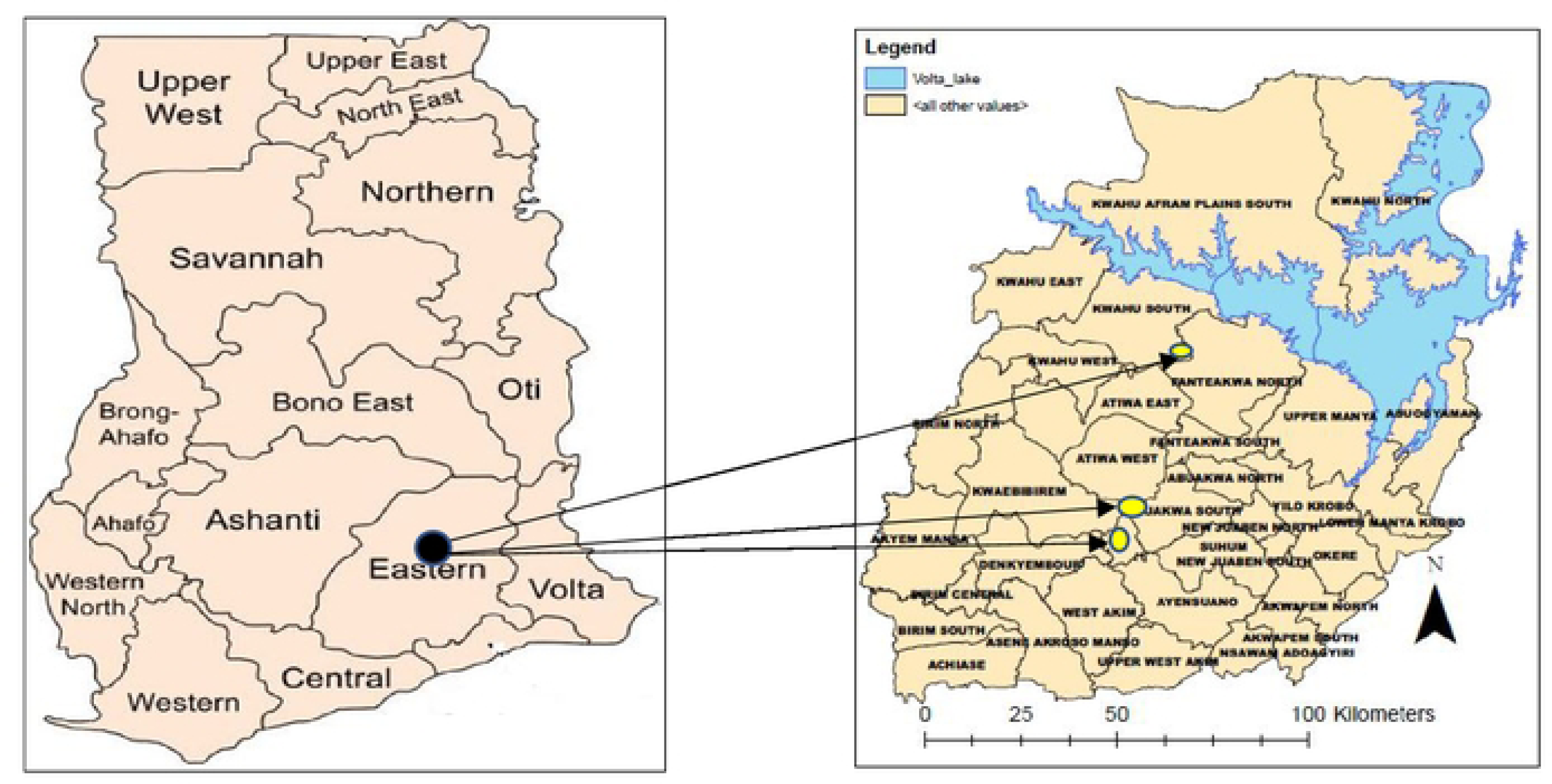
Map of Ghana with the 16 administrative regions Fig 2: Map of Eastern Region showing the three study sites.

The study employed a census among the health care workers who were actively involved in the operation of the IDSR system at the health facilities. Hence only the health staff who were actively involved in the routine disease surveillance activities at the various health facilities were enrolled in the study. A total of 347 health care workers of varied cadre at hospitals, health centres, Community Based Health Planning and Services (CHPS) compounds, maternity homes and clinics were enrolled in the study. To be included in the study, the individual must be a healthcare worker for more than six months, must be working in the New Juaben South Municipality, Abuakwa South Municipality, and Fanteakwa North District for more than six months, must be actively involved in routine surveillance activities at the various health facility levels and he or she must consent to be part of the study. An individual was excluded from this study if he or she was not at post during the period of the study and does not consent to be part of the study.

### Design of the study

A descriptive-cross sectional design was used in this study. The study employed both quantitative and qualitative methods using structured questionnaires to extract data from the health facilities.

### Study population

The study population involved health care providers who are actively involved in the routine disease surveillance activities working at health facilities in the three districts at the time of the study.

### Data collection

Participants were enrolled and data were collected between January10th 2022, and March 30th, 2022 using structured questionnaire, and observation checklist. The structured questionnaire was developed and administered using Open Data Kit (ODK) software application. A questionnaire was used to obtain information on socio-demographic, core surveillance functions, data collection tools and logistics. A descriptive questionnaire was used to obtain information on socio-demographic, core surveillance functions, data collection tools and logistics, on IDSR system. Using the structured questionnaire, the quantitative method was used to collect information from the respondents in a form of counts outcomes and analyzed accordingly. A quantitative method was used to obtain information such as facility type, duration at current position, educational qualification among others. Direct observations were also carried out to verify the availability of logistics, data collection tools, case definitions, IDSR technical guidelines as well as evidence of routine data analysis and displayed graphs and charts. There was a review of secondary data for the weekly and monthly IDSR reports. The questionnaire is attached as Annex 1 in S1 Text

### Quality control

Quality control was ensured by carrying out a pre-test of the data collection tool in the New Juaben North Municipality to correct any misunderstanding of the questions and also ensure consistency of the findings. The records review involved the data generated by the IDSR reporting period for 2021. All the participants were duly informed about the study. All the completed questionnaires on the Open Data Kit (ODK) were cross-checked in the presence of the respondent to ensure that they were completed to reduce the number of missing data. Data extracted from the ODK was verified and cleaned to minimize errors.

### Data Analysis

Data on the Open Data Kit (ODK) was extracted in an excel format where the necessary data cleaning and corrections were carried out. The cleaned data on the excel sheet was imported into STATA version 16.1 for statistical analysis. Descriptive statistics such as frequency, mean, and standard deviation were used to analyse the socio-demographic characteristics. Subsequently, chi-square and binary logistic regression analyses were conducted to assess the association between the socio-demographic factors and the effectiveness of the IDSR at the facility level using the WHO protocol for the assessments of national communicable disease surveillance and response at the health facility level, together with core surveillance functions. The analysis was carried out in two stages. The first analysis was performed using chi-square analysis to determine the level of association of socio-demographic and IDSR related factors was followed by a binary logistic regression analysis between the IDSR factors and its effectiveness at 0.05 significance and 95% confidence interval.

### Description of the matrix for classifying health facilities

To determine the effectiveness of the IDSR at the health facility level, a tool was adopted from the WHO protocol for the assessments of national communicable disease surveillance and response together with core surveillance functions (detection, reporting, analysis, confirmation, preparedness, response and feedback) for the disease surveillance system as described in the second edition IDSR technical guidelines [10], [11]. The responses obtained from the staff interviewed in the respective health facilities were used to determine whether staff were operating an effective disease surveillance system or not. A total of seven (7) indicators adopted from the WHO protocol for the assessment of national communicable disease surveillance and response together with the core surveillance functions were used for the assessment. Each of the indicators was assigned a score of one (1). Surveillance systems meeting effective IDSR system should score cumulative score of seven (7) and a score of less than seven (7) was classified as not effective (Table 1). Table 1 shows the various indicator variables with their operational definitions.

**Table 1:** Matrix for scoring assessing IDSR system.

| **Core functions** | **Detection and diagnosis** | **Reporting** | **Analysis** | **Investigation/ Specimen collection** | **Preparedness** | **Response based on data** | **Feedback** |
| --- | --- | --- | --- | --- | --- | --- | --- |
| **Definition** | Availability of case definitions at consulting rooms | Use of case definition to detect priority diseases | Availability of analyzed data on priority diseases | Investigation of priority disease (s) identified | Existence of response plans, logistics, referral systems etc | Evidence of response based on surveillance data | Dissemination of feedback to stakeholders |
| **Score** | Available=1<br>Not available=0 | Detection of priority disease for the past year =1<br>Inability to detect priority disease=0 | Availability of analyzed data=1<br>Data not analyzed =0 | Evidence of cases investigated=1<br>Priority disease not investigated=0 | Available=1<br>Not available=0 | Evidence of response=1<br>Non-existence of response action=0 | Evidence of feedback to stakeholders =1<br>Non-existence of evidence of feedback=0 |

**Note:** Facilities that scored seven (7) points were classified as effective and those that scored less than 7 were classified not effective (Table 1).

### Ethical approval

Written inform consent was obtained from the study participants. Ethical clearance for this research work was obtained from the Ghana Health Service Ethical Review Committee with an approved number GHS-ERC:041/10/21. Permissions were obtained from the New Juaben South, Abuakwa South, and Fanteakwa North Municipal/District Health Directorates, before the commencement of the study. All respondents who agreed to participate in the research work were well informed about the objective of the study. Participation in this study was voluntary, and respondents were not under any obligation to respond to questions or participate in the study if they do not want to do so.

## Results

### Socio-Demographic characteristics of the study participants

A total of three hundred and forty-seven (347) health care workers who are actively involved in the IDSR system participated in the study with the majority, (52.7 %,183/347) working at the Community Based Health Planning and Services (CHPS) compound. Most, (42.7% 148/347) of the study participants were aged 25-48 years (mean 31.3 years and SD ± 4.1 years) with the least (3.5%, 13/347) age bracket being 41-45 years. Females contributed to the highest proportion, (62.0%, 215/347) of the participants. Almost all (99.1%, 344/347) of the study participants had tertiary education. With regards to the number of years at current position of the health workers, (28.0%, 97/347) of them indicated they have worked for two (2) years at current position. Out of the 347 participants, 188(54.2%) were Community Health Nurses with the least number of cadre of staff, (1.2%, 4/347) being Medical Officers (Table 2).

**Table 2:** Socio-demographic characteristics of the study participants.

| **Variable** | **Frequency (n=347)** | **Percent (%)** |
| --- | --- | --- |
| **Facility type** |  |  |
| CHPS | 183 | 52.7 |
| Health Centre | 141 | 40.6 |
| Hospital | 15 | 4.3 |
| Clinic | 5 | 1.4 |
| Maternity Home | 3 | 0.9 |
| **Age groups** |  |  |
| 25-30 | 148 | 42.7 |
| 31-35 | 121 | 34.9 |
| 36-40 | 53 | 15.3 |
| 41-45 | 12 | 3.5 |
| 45 and above | 13 | 3.8 |
| **Sex** |  |  |
| Male | 132 | 38.0 |
| Female | 215 | 62.0 |
| **Education Qualification** |  |  |
| Tertiary | 344 | 99.1 |
| Secondary Education | 3 | 0.9 |
| **Duration at current position(years)** |  |  |
| <1 | 67 | 19.3 |
| 1 year | 64 | 18.4 |
| 2 years | 97 | 28.0 |
| 3 years | 26 | 7.5 |
| > 3 years | 93 | 26.8 |
| **Cadre of staff** |  |  |
| Community Health Nurse | 188 | 54.2 |
| Disease Control Officer | 55 | 15.9 |
| Enrolled Nurse | 50 | 14.4 |
| Midwife | 21 | 6.1 |
| Physician Assistant | 20 | 5.8 |
| Records Officer | 9 | 2.6 |
| Medical Officer | 4 | 1.2 |

With regards to the availability of rumor register and IDSR technical guidelines at health facilities, most of them (56.2%, 195/347; 69.7%, 242/347 respectively) indicated that they were available. Majority, (95.7%, 332/347) of the health care workers mentioned that they have immediate reporting forms for reporting priority diseases (90.8%, 315/347) of the respondents had IDSR weekly reporting forms available at their facilities while, 93.7% (325/347) of the participants had monthly IDSR reporting forms available for reporting. More than half, 194 (54.9%, 94/347) of the study participants had either a laptop or computer for managing surveillance data. More than half, (55.6%, 193/347) of the participants indicated that specimen carriers for sample transportation were available. Most (64.8%, 225/347) of the respondents had means of transport for disease surveillance activities. Almost all (97.1%, 337/347) of the health care workers had standard case definition at their facilities (Table 3). It was observed that most (82.7%, 287/347) of the respondents do conduct records review. More than half (54.47%, 189/347) of the health care workers have the capacity to apply case detection to detect diseases. About half (51.9%, 180/347) of the participants revealed that they did not receive any feedback from the next higher level in the past year while (42.9%,149/347) of the respondents indicated that they did not have surveillance meeting with their communities in the previous year (Table 3).

**Table 3:**
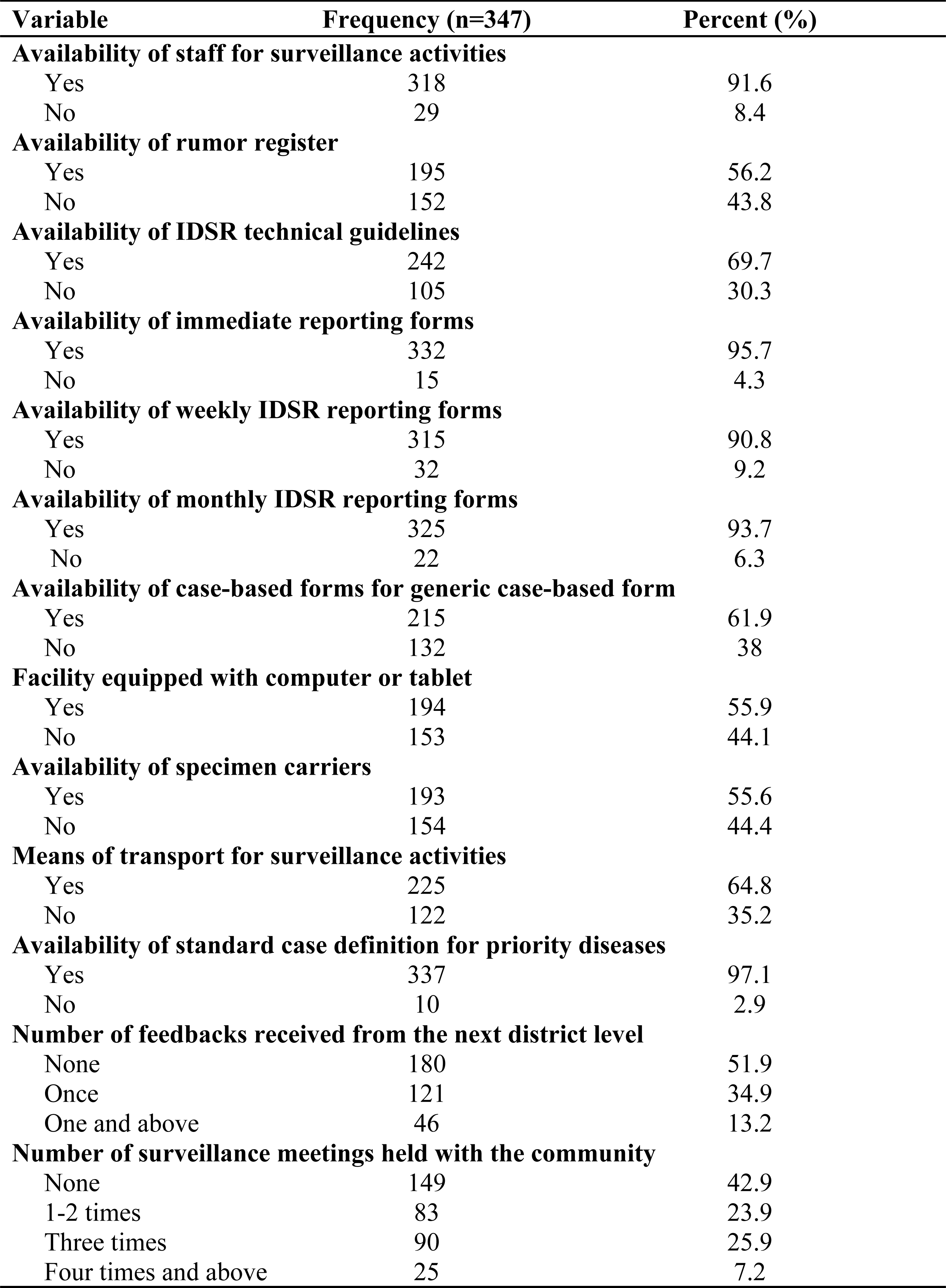
Integrated Disease Surveillance and Response related factors Variable Frequency (n=347) Percent (%)

| **Variable** | **Frequency (n=347)** | **Percent (%)** |
| --- | --- | --- |
| **Availability of staff for surveillance activities** |  |  |
| Yes | 318 | 91.6 |
| No | 29 | 8.4 |
| **Availability of rumor register** |  |  |
| Yes | 195 | 56.2 |
| No | 152 | 43.8 |
| **Availability of IDSR technical guidelines** |  |  |
| Yes | 242 | 69.7 |
| No | 105 | 30.3 |
| **Availability of immediate reporting forms** |  |  |
| Yes | 332 | 95.7 |
| No | 15 | 4.3 |
| **Availability of weekly IDSR reporting forms** |  |  |
| Yes | 315 | 90.8 |
| No | 32 | 9.2 |
| **Availability of monthly IDSR reporting forms** |  |  |
| Yes | 325 | 93.7 |
| No | 22 | 6.3 |
| **Availability of case-based forms for generic case-based form** |  |  |
| Yes | 215 | 61.9 |
| No | 132 | 38 |
| **Facility equipped with computer or tablet** |  |  |
| Yes | 194 | 55.9 |
| No | 153 | 44.1 |
| **Availability of specimen carriers** |  |  |
| Yes | 193 | 55.6 |
| No | 154 | 44.4 |
| **Means of transport for surveillance activities** |  |  |
| Yes | 225 | 64.8 |
| No | 122 | 35.2 |
| **Availability of standard case definition for priority diseases** |  |  |
| Yes | 337 | 97.1 |
| No | 10 | 2.9 |
| **Number of feedbacks received from the next district level** |  |  |
| None | 180 | 51.9 |
| Once | 121 | 34.9 |
| One and above | 46 | 13.2 |
| **Number of surveillance meetings held with the community** |  |  |
| None | 149 | 42.9 |
| 1-2 times | 83 | 23.9 |
| Three times | 90 | 25.9 |
| Four times and above | 25 | 7.2 |

### Socio-demographic factors associated with IDSR system

There was significant association between the sex of the participants (Chi2=6.32, P<0.05) and number of years at current position (Chi2=11.52, P<0.05) and the effectiveness of the IDSR system. Meanwhile in the regression model and after adjusting for multiple factors, respondents who had served for more than three years at their current position were about three times more likely to have an effective surveillance system in place compared to respondents who had served less than a year at their current position (AOR= 2.94; 95% CI= 1.15-7.52; p= 0.02) (Table 4).

**Table 4:**
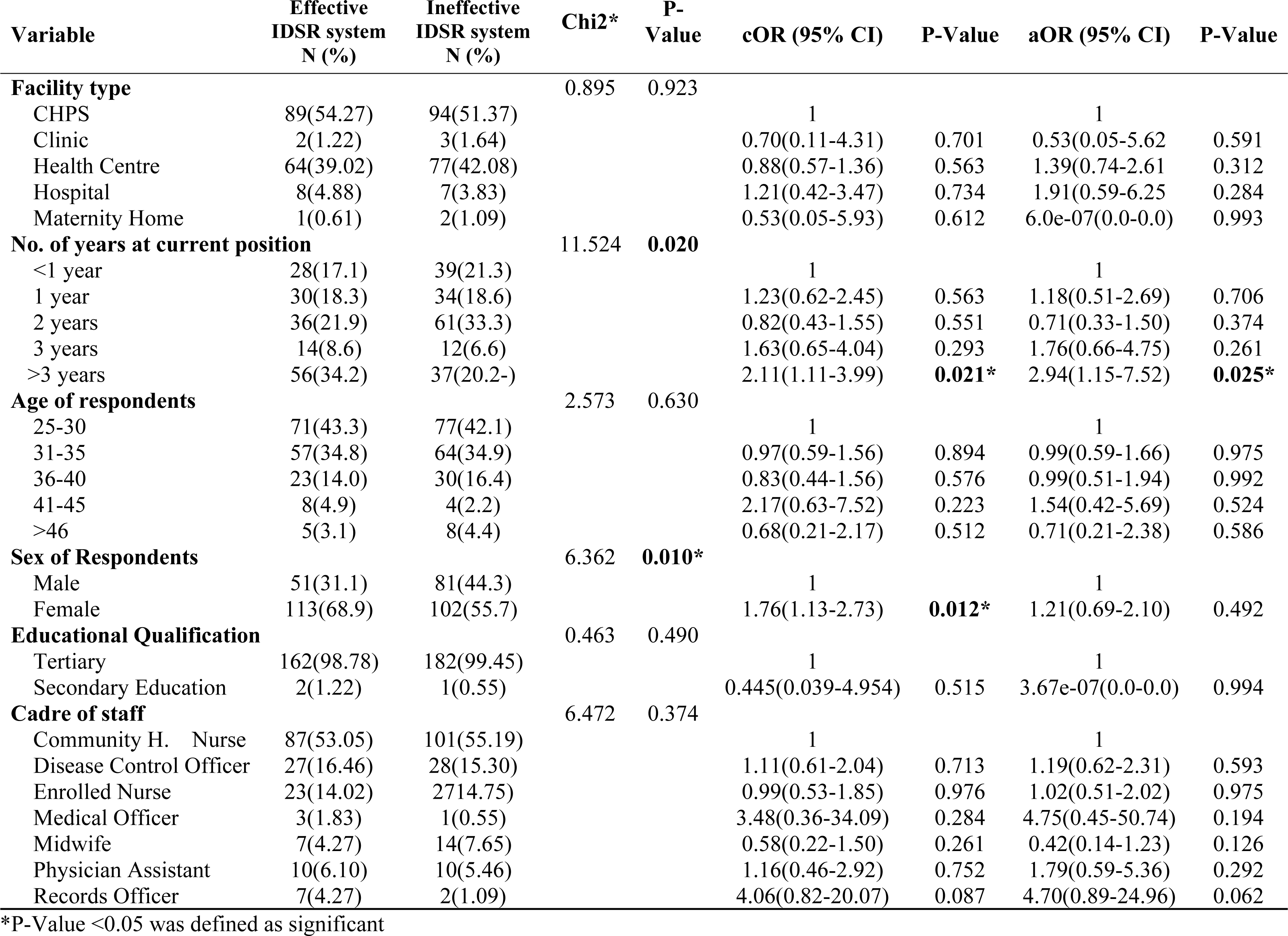
Socio-demographic factors associated with IDSR system.

### Data collection tools related factors associated with IDSR system

Table 5 displays the factors associated with data collection tools and the effectiveness of the IDSR system. There was statistically significant association between the availability of rumor register and the effectiveness of the IDSR system (Chi2=10.34, P<0.001). There was also statistically significant association between the availability of immediate reporting forms and the effectiveness of the Integrated Disease Surveillance and Response system (Chi2=4.67, P<0.05). The association between availability of COVID-19 case based reporting forms and the effectiveness of the Integrated Disease Surveillance and Response system showed strong association (Chi2=6.73, P<0.05). No significant associations were found between AFP, measles, yellow fever and meningitis case-based forms as well as weekly and monthly IDSR reporting forms.

**Table 5:** Data collection tools related factors associated with IDSR system.

| Variable | Effective<br>IDSR<br>system<br>N (%) | Ineffective<br>IDSR<br>system<br>N (%) | Chi2* | P-Value | cOR (95% CI) | P-Value | aOR (95% CI) | P-Value |
| --- | --- | --- | --- | --- | --- | --- | --- | --- |
| **Availability of rumor register** |  |  | 10.34 | **<0.001\*** |  | **0.001\*** |  | **0.011\*** |
| Yes | 107(65.2) | 88(48.1) |  |  | 1 |  | 1 |  |
| No | 57(34.8) | 95(51.9) |  |  | 2.03(1.31-3.12) |  | 1.97(1.15-3.37) |  |
| **Availability of immediate reporting forms** |  |  | 4.67 | **0.03\*** |  | **0.042\*** |  | **0.001\*** |
| Yes | 161(98.2) | 171(93.4) |  |  | 1 |  | 1 |  |
| No | 3(1.8) | 12(6.6) |  |  | 3.77(1.04-13.59) |  | 12.67(2.74-58.6) |  |
| **Availability of weekly IDSR reporting forms** |  |  | 0.113 | 0.741 |  | 0.752 |  | 0.894 |
| Yes | 148(90.2) | 167(91.3) |  |  | 1 |  |  |  |
| No | 32(9.8) | 16(8.7) |  |  | 0.88(0.42-1.83) |  | 0.94(0.43-2.05) |  |
| **Availability of monthly IDSR reporting forms** |  |  | 0.036 | 0.860 |  | 0.863 |  | 0.771 |
| Yes | 154(93.9) | 171(9.3) |  |  | 1 |  | 1 |  |
| No | 22(6.1) | 12(6.6) |  |  | 1.08(0.45-2.57) |  | 0.86(0.33-2.24) |  |
| **Availability of AFP case-based forms** |  |  | 2.244 | 0.132 |  | 0.141 |  | 0.184 |
| Yes | 157(95.7) | 168(91.8) |  |  | 1 |  | 1 |  |
| No | 7(4.3) | 15(8.2) |  |  | 2.0(0.79-5.04) |  | 2.13(0.70-6.52) |  |
| **Availability of measles case-based forms** |  |  | 0.387 | 0.536 |  | 0.532 |  | 0.723 |
| Yes | 155(94.0) | 170(92.9) |  |  | 1 |  | 1 |  |
| No | 22(6.0) | 13(7.1) |  |  | 1.31(0.54-3.16) |  | 1.24(0.37-4.07) |  |
| **Availability of meningitis case-based forms** |  |  | 2.123 | 0.154 |  | 0.144 |  | 0.871 |
| Yes | 149(90.9) | 157(85.8) |  |  | 1 |  | 1 |  |
| No | 15(9.1) | 26(14.2) |  |  | 1.64(0.83-3.22) |  | 0.91(0.32-2.61) |  |
| **Availability of yellow fever case-based forms** |  |  | 2.831 | 0.091 |  | 0.091 |  | 0.264 |
| Yes | 153(93.3) | 161(87.9) |  |  | 1 |  | 1 |  |
| No | 11(6.7) | 22(12.1) |  |  | 1.90(0.89-4.05) |  | 2.05(0.58-7.21) |  |
| **Availability of COVID-19 case based forms** |  |  | 6.733 | **0.012\*** |  | **0.012\*** |  | **0.034\*** |
| Yes | 146(89.1) | 144(78.7) |  |  | 1 |  | 1 |  |
| No | 18(10.9) | 39(21.3) |  |  | 2.19(1.20-4.01) |  | 3.01(1.06-8.56) |  |
| **Availability of generic case-based forms for priority diseases** |  |  | 4.223 | **0.032\*** |  | **0.013\*** |  | 0.734 |
| Yes | 145(89.5) | 142(76.8) |  |  | 1 |  | 1 |  |
| No | 17(10.5) | 43(23.2) |  |  | 1.75(1.13-2.73) |  | 0.88(0.44-1.76) |  |

\*P<0.05 was defined as significant

After adjusting for possible confounding effect of the variables in the regression model, the availability of rumor register at health facilities for documentation and investigating rumors is about 2 times more likely to contribute to an effective disease surveillance system compared to facilities without rumor registers (AOR= 1.97; 95% CI= 1.15-3.37; p<0.05). Adjusting for generic case-based forms for priority diseases, monthly and weekly IDSR reporting forms, availability of COVID-19 case based forms, AFP, measles, meningitis, and yellow fever case based, the availability of immediate case reporting forms was about 12.7 times more likely to contribute to the effectiveness of IDSR system at health facilities compare to facilities without immediate reporting forms (AOR=12.67; 95% CI=2.74-58.62; P<0.001). The availability of COVID-19 case investigation form at heath facilities was 3 times more likely to contribute to an effective IDSR system at health facilities after adjusting for the effect of other variables and confounders (AOR=3.01; 95 CI=1.06-8.56; P<0.05) (Table 5).

### Logistics and case detection related factors associated with IDSR system

The means of transport for surveillance activities was statistically significantly (Chi2=8.12, P<0.001) associated with the effectiveness of the IDSR system. No significant association was observed between availability of staff for surveillance activities, facility equipped with either a computer or a laptop for data management, availability of specimen carriers, and availability of sample containers for AFP, blood, cerebrospinal fluid, and sputum samples. After adjusting for possible confounding factors in the regression model, the availability of transport for IDSR activities was almost 3.4 times more likely to contribute to an effective disease surveillance system compared to health facilities without available means of transportation for disease surveillance activities (AOR= 3.36; 95% CI= 1.44-7.83; p=0.00 (Table 6).

**Table 6:** Case detection and logistics related factors associated with IDSR system.

| Variable | Effective<br>IDSR<br>system<br>N (%) | Ineffective<br>IDSR<br>system<br>N (%) | Chi2* | P-Value | cOR (95% CI) | P-Value | aOR (95% CI) | P-Value |
| --- | --- | --- | --- | --- | --- | --- | --- | --- |
| **Facility equipped with computer or tablet for data** |  |  | 1.864 | 0.174 |  | 0.171 |  | 0.544 |
| Yes | 98(59.8) | 96(52.5) |  |  | 1 |  | 1 |  |
| No | 66(40.2) | 87(47.5) |  |  | 1.34(0.87-2.06) |  | 0.81(0.41-1.59) |  |
| **Availability of specimen carriers for** |  |  | 2.835 | 0.093 |  | 0.093 |  | 0.321 |
| Yes | 99(60.4) | 94(51.4) |  |  | 1 |  | 1 |  |
| No | 65(39.6) | 89(48.6) |  |  | 1.44(0.94-2.21) |  | 1.50(0.67-3.35) |  |
| **Availability of staff for surveillance activities** |  |  | 0.793 | 0.73 |  | 0.373 |  | 0.187 |
| Yes | 148(90.2) | 170(92.9) |  |  | 1 |  |  |  |
| No | 16(9.8) | 13(7.10) |  |  | 0.70(0.32-1.51) |  | 2.08(0.70-6.14) |  |
| **Means of transport for surveillance activities** |  |  | 8.123 | **0.001\*** |  | **0.005\*** |  | **0.005\*** |
| Yes | 119(72.6) | 106(57.9) |  |  | 1 |  | 1 |  |
| No | 45(27.4) | 77(42.1) |  |  | 1.92(1.22-3.01) |  | 3.36(1.44-7.83) |  |
| **Availability of consulting room register** |  |  | 0.485 | 0.487 | 0.486 |  |  | 0.274 |
| Yes | 144(87.8) | 156(85.3) |  |  | 1 |  | 1 |  |
| No | 20(12.2) | 27(14.8) |  |  | 1.24(0.66-2.31) |  | 0.58(0.22-1.53) |  |
| **Conduct records review** |  |  | 4.382 | **0.036\*** |  | **0.035\*** |  | 0.194 |
| Yes | 143(87.2) | 144(78.7) |  |  | 1 |  | 1 |  |
| No | 21(12.8) | 39(21.3) |  |  | 1.88(1.03-3.28) |  | 1.71(0.76-3.81) |  |
| **Ability to apply case definition for case detection** |  |  | 18.053 | **<0.001\*** |  | **<0.001\*** |  | **0.013\*** |
| Yes | 109(66.5) | 80(43.7) |  |  | 1 |  | 1 |  |
| No | 55(33.5) | 103(56.3) |  |  | 2.55(1.64-3.94) |  | 1.94(1.17-3.21) |  |

\*P<0.05 was defined as significant

There was a significant association between reviewing of records and the effectiveness of the IDSR system in the chi square analysis (Chi2= 4.36, P<0.05). The ability of the study participants to apply case definition in case detection was also significantly associated with the effectiveness of the IDSR system (Chi2=18.05, P<0.001). Availability of consulting room register at health facilities was not found to be associated with the effectiveness of the IDSR system. In the regression model, respondents who have the capacity to apply case definition for case detection were approximately 2 times more likely to have an effective disease surveillance system compared to health workers who cannot apply case definition for detecting cases (AOR= 1.94; 95% CI= 1.17-3.21; p=0.013) after adjusting for possible confounding factors (Table 6).

### Disease reporting related factors associated with IDSR system

Table 7 summarizes the disease reporting related factors associated with an effective surveillance system. There was a statistically significant association between means of reporting surveillance information to the next level and the effectiveness of the IDSR system (Chi2=27.27, P<0.001). There was no significant association between completeness of weekly and monthly reporting, and the lack of these forms for the last six (6) months and the effectiveness of the IDSR system. After adjusting for possible confounding factors, health care workers who reported to the next level using electronic or paper-based were 55% (AOR= 0.45; 95% CI= 0.25-0.80; p= 0.007) and 81% (AOR= 0.19; 95% CI= 0.09-0.39; p<0.001) respectively less likely to have an effective surveillance system compared to respondents who used both means of reporting to the next level (Table 7).

**Table 7:**
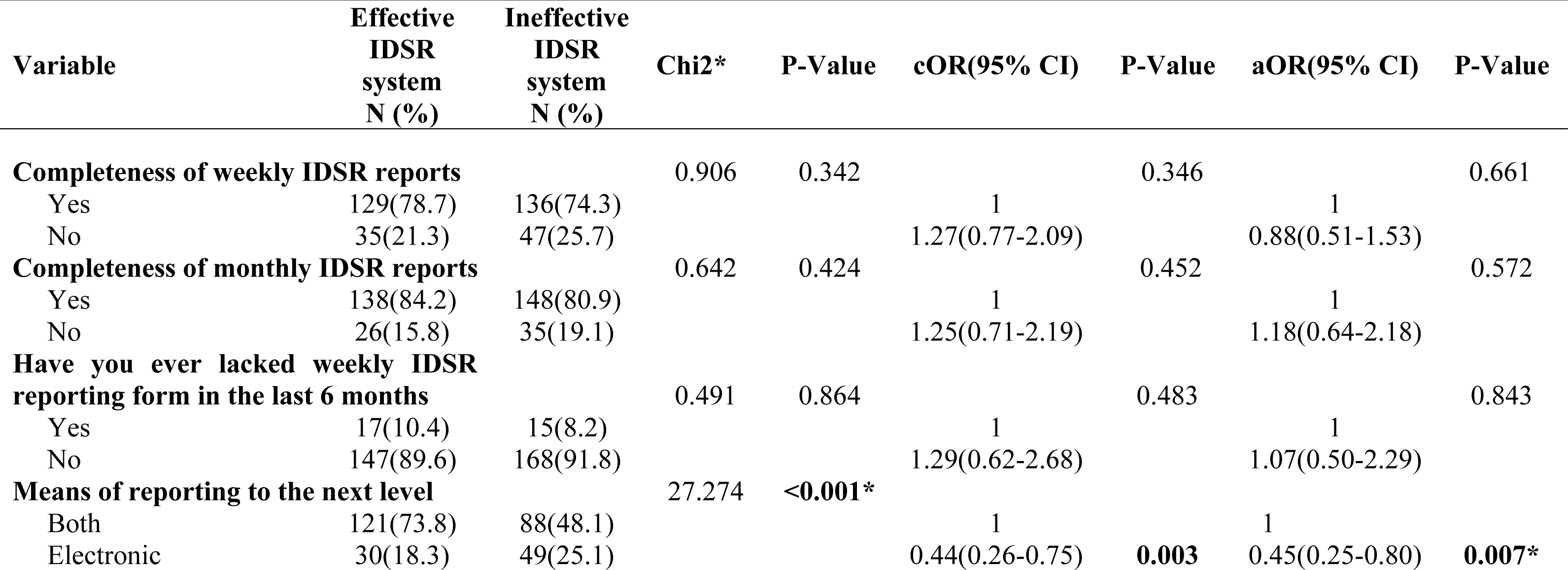
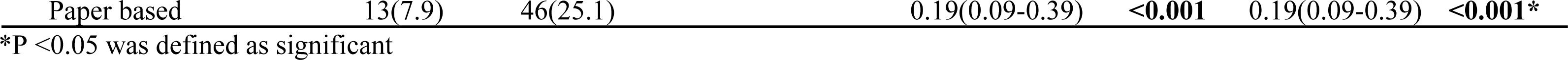
Disease reporting related factors associated with IDSR system.

### Disease confirmation related factors associated with IDSR system

The capacity of the health facility to handle specimen before shipment or transportation was statistically significantly associated with the effectiveness of the IDSR system (Chi2= 20.86, P<0.001). There was also a significant association between the effectiveness of IDSR system and the availability of packaging materials for shipment (Chi2= 12.302, P<0.001) and the capacity of the facility to process samples for laboratory investigation (Chi2= 5.20, P=0.023). After adjusting for possible confounding effect of the variables, the odds of contributing to effective Integrated Disease Surveillance and Response among health facilities that have the capacity to handle samples before shipment or transportation was 3.3 times higher compared to facilities without the capacity to handle samples before shipment or transport for investigation (AOR=3.34; 95 CI=0.57-3.04; P=0.006) (Table 8).

**Table 8:**
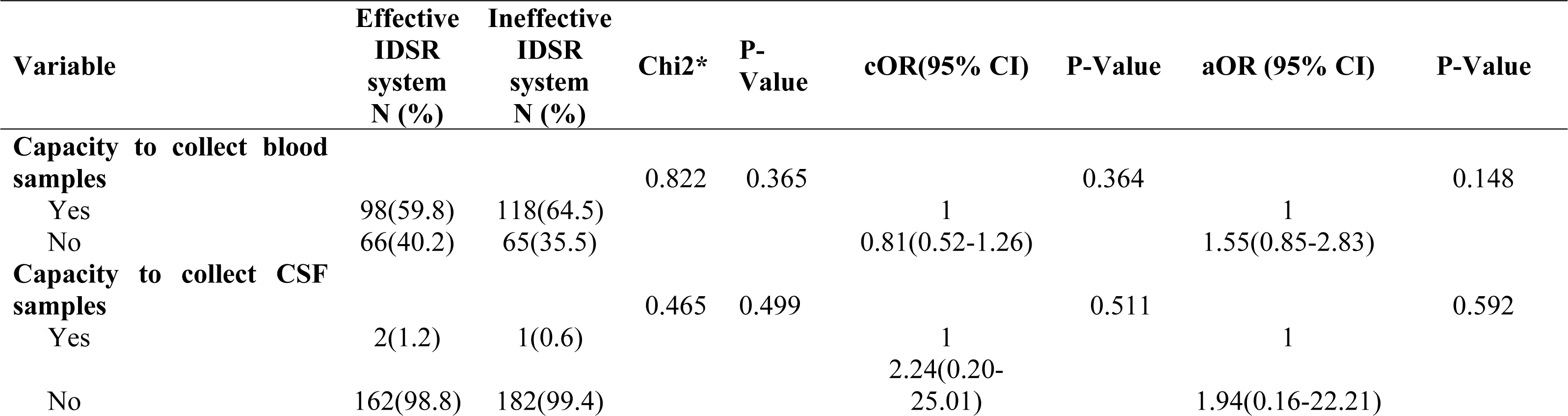
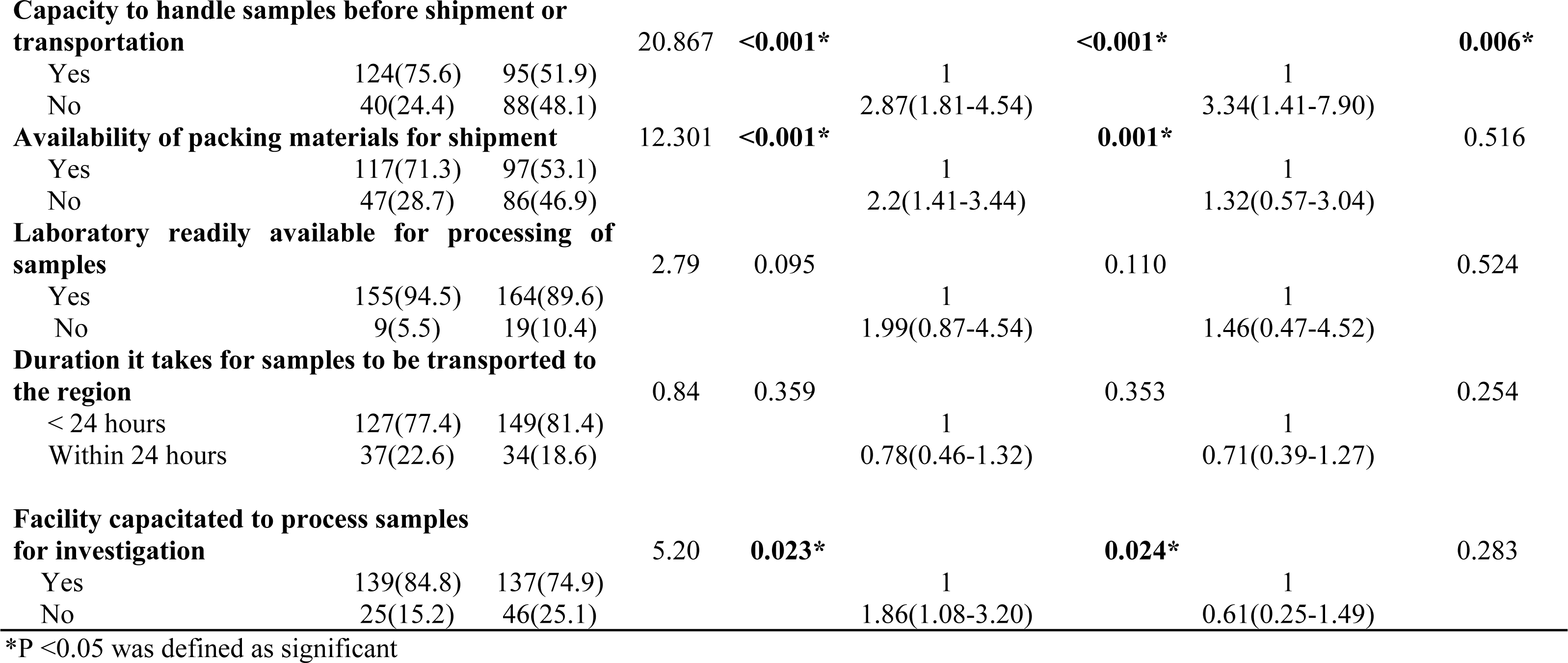
Disease confirmation related factors associated with IDSR system.

### Disease Response and feedback related factors associated with IDSR system

The availability of threshold for epidemic prone diseases was found to be significantly associated with the effectiveness of the IDSR system (Chi2=8.552, P<0.05). There was also a statistically significant association between resources availability to contain disease outbreaks and the effectiveness of the IDSR system (Chi2=50.319, P<0.001). The duration it takes for the next higher level to respond to surveillance needs, existence of rapid response team and availability of minutes for rapid response meetings did not show any significant association with the effectiveness of the IDSR system.

There was significant association between receipt of feedback from the next higher level and the effectiveness of the IDSR system (Chi2=10.59, P<0.001). The number of meetings on disease surveillance held with the community also showed statistically significant relationship with the effectiveness of the IDSR system (Chi2=19.48, P<0.001). After adjusting for possible confounding variables, respondents who did not receive feedback from the next higher level were 52% less likely to have an effective surveillance system compared to respondents who received feedback from the next higher level (AOR= 0.48; 95% CI= 0.23-1.00; p= 0.05). Also, respondents who held meetings on surveillance related activities with the community 3 times were about 5 (AOR= 5.21; 95% CI= 2.43-11.18; p<0.001) times more likely to have an effective surveillance system compared to respondents who held 1-2 meetings on surveillance activities with the TABLEREF (Table 9).

**Table 9:** Disease response and feedback related factors associated with IDSR system.

| Variable | Effective<br>IDSR<br>system<br>N (%) | Ineffective<br>IDSR<br>system<br>N (%) | Chi2* | P-Value | cOR(95% CI) | P-Value | aOR(95% CI) | P-Value |
| --- | --- | --- | --- | --- | --- | --- | --- | --- |
| **Duration it takes for next higher level to respond to surveillance needs** |  |  | 0.3292 | 0.848 |  |  |  |  |
| 72 hours and above | 6(3.67) | 7(3.8) |  |  | 1 |  | 1 |  |
| Within 24 hours | 114(69.5) | 122(66.7) |  |  | 1.09(0.35-3.34) | 0.882 | 1.59(0.50-5.07) | 0.426 |
| Within 48 hours | 44(26.8) | 54(29.5) |  |  | 0.95(0.28-3.03) | 0.934 | 2.77(0.78-9.84) | 0.113 |
| **Availability of thresholds for epidemic prone diseases** |  |  | 8.552 | **0.003\*** |  | 0.185 |  | 0.215 |
| Yes | 113(68.9) | 98(53.6) |  |  | 1 |  | 1 |  |
| No | 51(31.1) | 85(46.4) |  |  | 1.32(0.86-2.02) |  | 0.70(0.40-1.21) |  |
| **Availability of rapid response team** |  |  | 2.419 | 0.12 |  | 0.122 |  | 0.832 |
| Yes | 116(70.7) | 115(62.8) |  |  | 1 |  | 1 |  |
| No | 48(29.3) | 68(37.2) |  |  | 1.42(0.91-2.24) |  | 0.92(0.41-2.10) |  |
| **Evidence of minutes for rapid response** |  |  | 1.419 | 0.233 |  | 0.234 |  | 0.243 |
| Yes | 100(60.9) | 100(54.6) |  |  | 1 |  | 1 |  |
| No | 64(39.1) | 83(45.4) |  |  | 1.29(0.84-1.98) |  | 1.56(0.73-3.31) |  |
| **Facility resourced to contain outbreaks** |  |  | 50.319 | **<0.001\*** |  | **<0.001\*** |  | **<0.001\*** |
| Yes | 147(89.6) | 101(55.2) |  |  | 1 |  | 1 |  |
| No | 17(10.4) | 82(44.8) |  |  | 7.02(3.92-12.54) |  | 9.27(4.81-17.83) |  |
| **Receipt of feedback from the next higher level** |  |  | 10.594 | **0.001\*** |  |  |  | **0.054\*** |
| Yes | 45(100.0) | 243(80.5) |  |  | 1 |  | 1 |  |
| No | 0(0.0) | 59(19.5) |  |  | 1.07(0.61-1.88) | 0.821 | 0.48(0.23-1.00) |  |
| **Number of feedbacks received from the district for the previous year** |  |  | 4.362 | 0.112 |  |  |  |  |
| Once | 80(48.78) | 100(54.64) |  |  | 1 |  | 1 |  |
| 2 times | 66(40.24) | 55(30.05) |  |  | 1.50(0.94-2.38) | 0.082 | 1.35(0.76-2.37) | 0.294 |
| 3 times and above | 18(10.98) | 28(15.30) |  |  | 0.80(0.41-1.55) | 0.527 | 1.06(0.52-2.17) | 0.865 |
| **Number of meetings on surveillance held with community** |  |  | 19.483 | <0.001* |  |  |  |  |
| 1-2 times | 53(32.3) | 96(52.5) |  |  | 1 |  | 1 |  |
| 3 times | 54(32.9) | 29(15.9) |  |  | 3.37(1.92-5.91) | <0.001* | 5.21(2.43-11.18) | <0.001* |
| 4 times | 43(26.2) | 47(25.7) |  |  | 1.65(0.97-2.82) | 0.061* | 2.76(1.34-5.67) | 0.006* |
| None | 14(8.5) | 11(6.1) |  |  | 2.30(0.97-5.43) | 0.056* | 2.57(0.96-6.85) | 0.051* |
| **Discussion of surveillance related issues at review meetings** |  |  | 0.523 | 0.471 |  |  |  | 0.192 |
| Yes | 117(71.3) | 124(67.7) |  |  | 1 |  | 1 |  |
| No | 47(28.7) | 59(32.2) |  |  | 1.18(0.74-1.87) | 0.473 | 0.61(0.28-1.29) |  |

\*P <0.05 was defined as significant

## Discussion

This study demonstrated that the number of years at current position can significantly influence the IDSR System and this corroborates with a study conducted in Kenya which reported that the number of years of service has influence on the disease surveillance system and respondents who have served for more years are likely to have an effective surveillance system in place. This means that health facilities with health care workers who have worked for more yeas would have better disease surveillance system in place compare to health facilities with staff who have worked for less number of years in the field of disease surveillance [5].

There was significant association between the availability of rumor register and the effectiveness of the IDSR system. There was availability of rumor registers and IDSR technical guidelines in most of the health facilities which is consistent with other studies in Nigeria and Sierra Leone [12], [13] respectively where it was observed that IDSR technical guidelines, consulting room registers, rumor log book, data collection tools and case definitions were available in most of their health facilities. On the contrary, findings from a study in India indicated that there were no rumor log books for documenting rumors in all the health facilities [14]. The existence of rumor registers help in effective documentation of all rumors regarding disease outbreaks and other public health events so that follow ups and investigations can be carried in order to timely follow up and investigate possible outbreaks and events. Also, this study indicated that IDSR weekly and monthly reporting forms were available at their facilities for service delivery as opposed to the findings in Kenya where only 9.2% of health facilities were in short supply of weekly IDSR reporting forms [15]. The availability of weekly and monthly reporting forms help in effective documentation of priority diseases that are recorded on weekly and monthly basis so that thresholds can be analyzed for action where necessary.

Furthermore, interactions with the health care providers in this study revealed that more than half of the study participants had either a laptop or computer for managing surveillance data. This is different from the observation made in primary health care facilities in Oyo state, Nigeria where inadequate computers and reporting forms were some of the major challenges affecting the disease surveillance system [16]. An ideal disease surveillance system requires routine data collection, analysis, interpretation and feedback and that can best be done with the availability of computers and lap tops which means health care workers who have lap tops or computers are in the better position to operate the disease surveillance system effectively.

The availability of transport for IDSR activities was observed to be associated with effective disease surveillance system compared to health facilities without means of transportation for disease surveillance activities in this study. It was observed that more than half of the health care workers said they have means of transport for disease surveillance activities which agrees with a study in Liberia which reported that there was availability of motorbikes for disease surveillance activities in almost all the facilities which participated in the study [17]. Similary, the findings from a study, in Southern Ethiopia, found that bicycles, motorcycles and vehicles were accessible for routine surveillance activities [18]. In this study, availability of IDSR technical guidelines, availability of specimen carriers and availability of sample containers did not influence the IDSR system. The IDSR technical guidelines serve as reference material that contains the necessary protocols for the operation of the disease surveillance system. The investigation of most priority disease require the use of specimen containers and sample carriers to help in the sample transportation processes. Findings from this study revealed that almost all the health care workers had standard case definition at their facilities serving as a guide in the detection and investigation of priority diseases which contradicts with the observations made in Yemen [19] which reported that most of the health facilities assessed do not have case definitions at their disposal and this affects effective diagnosis and case detection [19]. Review of records helps in the early detection and investigation of priority diseases that may be missed by clinicians. This study has demonstrated that most of the health care workers do conduct records review routinely in their various health facilities which corroborates with the findings in a Kenyan study which reported that standard case definitions for IDSR were available in all health facilities to aid in records review and case detection [20].

Majority of the health care workers who participated in this study indicated that they have not ever lacked weekly IDSR reporting form, which is in line with a research carried out in northern Ghana which found that over 75% (34/47) of the informants said routine disease surveillance data reporting was good because of the availability of weekly and monthly IDSR reporting forms on a regular basis [21]. So this practice needs to be maintained and encouraged to ensure effective monitoring of pathogens that have the potential to cause epidemics or pandemics.

The use of both electronic and paper-based reporting give room for all levels of officers regardless of their knowledge level on the use of computer to operate the IDSR system to achieve its objective. In this current study, most of the respondents indicated that they use both paper-based and electronic means of reporting which is in contrast with other studies which reported that only 70% (33/47) of the countries were practicing electronic IDSR system in their service delivery activities [3].

Findings from this current study revealed that weekly and monthly IDSR report completeness and timeliness were about 80.0% similar to other studies [22] and can contribute to an effective disease surveillance system. This study observed that majority of the respondents had analyzed their surveillance data by person, place and time and this is supported by other studies in Ghana and Liberia [17], [21]. The reason for the high rate of analyzed data could be attributed to the fact that the facilities require these analyzed data for feedback dissemination with relevant stakeholders. In addition, a research conducted in Northern Ghana [21] reported that about half of the participants analyze their routine surveillance data but the reports are not posted on their notice boards. Contrary to the findings of this study and some others, conducted in Ethiopia and Tanzania respectively, they found that lack of capacity to conduct data analysis was the major reason for the inadequate data analysis in most of the health facilities [18], [23].

The availability of packaging materials was found to contribute to an effective disease surveillance system. The availability of packaging materials help to ensure that samples are safely transported to the laboratory for investigation. It is also important to note that other studies revealed different observation where there was low capacity of health facilities to process samples for laboratory investigation due to inadequate capacity of staff and the necessary resources [20]–[22]. Timely submission of samples from the facility level to the regional level for laboratory investigation was observed in this study as reported by other studies [5], where samples were transported on time for laboratory investigation.

In this study more than half of the study participants had threshold for epidemic prone diseases which confirms the assertion that health care workers have the epidemic thresholds for priority diseases in a study conducted in Tanzania to assess the core and support functions of the IDSR system [24].The availability of threshold for epidemic prone diseases has influence on the IDSR system.

Feedback regarding disease surveillance activities help to prompt health care workers on the performance and shortfalls regarding routine operation of the disease surveillance activities across health facilities. One of the important findings from this study was that more than half of the health workers did not receive any feedback from the next higher level in the past year which is similar to other observations where there was inadequate feedback for IDSR system at the health systems and among stakeholders [20].The implications for lack of feedback in the routine operation of the IDSR is that it affects the optimal performance of the system in detection,, prevention and controlling of infectious diseases. In Yemen and India there were reports showed that there were inadequate feedback across the various levels of health administration [14], [19].

The limitation of the study was that the study was cross-sectional in nature and could not provide all relevant Disease Surveillance information for generalization to the entire disease surveillance system in the Eastern Region. Again, the study was limited to IDSR system at health facilities hence findings from the study cannot be used to assess the entire disease surveillance system in the three districts which participated in the study.

### Conclusions

The study observed that availability of rumor register at health facilities contributes to an effective Integrated Disease Surveillance system. Also, availability of immediate case reporting forms for investigating priority diseases is associated with the effectiveness of the disease surveillance system. Furthermore, means of transport for surveillance activities have positive influence on the IDSR system. The capacity of health care workers to apply case definition also contributes to the effectiveness of the IDSR system. Again, more than half of the health workers did not receive feedback from the next higher level in the past year. It is recommended that the District Health Directorates should ensure that feedback regarding surveillance activities from all levels are disseminated to the health facilities.

### Ethics approval and consent to participate

Ethical clearance for this research work was obtained from the Ghana Health Ethical Review Committee with an approved number GHS-ERC:041/10/21. Permissions were obtained from the New Juaben South, Abuakwa South, and Fanteakwa North Municipal/District Health Directorates, before the commencement of the study. Written informed consent was obtained from respondents. All respondents who agreed to participate in the research work were well informed about the aim of the study. Participation in this study was voluntary, and respondents were not under any obligation to respond to questions or participate in the study if they do not want to do so.

### Availability of data and materials

The data used in this study was sourced from health facilities through interviews. All data generated or analyzed for this study are available for official purposes at annex 1 (S1, S2 and S3).

## Data Availability

All relevant data are within the manuscript and its Supporting Information files

## Acknowledgements

The authors are thankful to the Ghana Health Service Ethical Review Committee. We are also grateful to the Eastern Regional Health Directorate for the support. Again, we appreciate the support of District Health Management Teams of the New Juaben South, Abuakwa South Health and Fanteakwa North and the health workers for participating in the study. We are also thankful to the Research Assistants who collected the data.

## Supporting information

**S1 Text. Annex 1 questionnaire.** (PDF)

**S2 XML. Designed Microsoft Excel Form which was used to develop the questionnaire on the ODK** (XLSX)

**S3 XML. Formatted data that was used to do the analysis on STATA** (XLSX)

## Author contributions

**Conceptualization:** Paul Twene, Bismark Sarfo, Alfred A.E Yawson, John Ekow Otoo, Annette Asraku

**Data curation:** Paul Twene

**Formal analysis:** Paul Twene, Bismark Sarfo

**Funding acquisition:** Paul Twene, Bismark Sarfo, John Ekow Otoo,

**Investigation:** Paul Twene

**Methodology:** Paul Twene, Bismark Sarfo, Alfred A.E Yawson

**Project administration:** Paul Twene, Bismark Sarfo, John Ekow Otoo, Annette Asraku

**Resources:** Paul Twene, Bismark Sarfo, John Ekow Otoo

**Software:** Paul Twene

**Supervision:** Bismark Sarfo, Alfred A.E Yawson, John Ekow Otoo

**Validation:** Paul Twene

**Writing** –**original draft:** Paul Twene

**Writing** –**review and editing:** Paul Twene, Bismark Sarfo, Alfred A.E Yawson, John Ekow Otoo, Annette Asraku

